# Memory-Dependent Model for the Dynamics of COVID-19 Pandemic

**DOI:** 10.1101/2020.06.26.20141242

**Authors:** K.M. Furati, I.O. Sarumi, A.Q.M. Khaliq

**Author notes:** Email addresses (K.M. Furati), (I.O. Sarumi), (A.Q.M. Khaliq).

## Abstract

COVID-19 pandemic has impacted people all across the world. As a result, there has been a collective effort to monitor, predict, and control the spread of this disease. Among this effort is the development of mathematical models that could capture accurately the available data and simulate closely the futuristic scenarios. In this paper, a fractional-order memory-dependent model for simulating the spread of COVID-19 is proposed. In this model, the impact of governmental action and public perception are incorporated as part of the time-varying transmission rate. The model simulation is performed using the two-step generalized exponential time-differencing method and tested for data from Wuhan, China. The mean-square errors demonstrate the merit of the fractional-order model and provide a good estimate of the optimal order.

## 1. Introduction

The novel coronavirus pandemic, COVID-19, has impacted all aspects of our life and the disease continues to threaten global economy, public health, human interactions, and safety. Since the outbreak of the disease, there has been a collective effort to monitor, predict, and control its spread. For this purpose, different biological and non-biological techniques, tools, interventions, analysis, and experiments have been employed to understand the dynamics of disease spread.

Epidemiological mathematical models continue to play a pivotal role in understanding, capturing and predicting the spread of infectious diseases. In particular, first-order models such as SIR, SEIR, and many others, are intensively explored to mimic the collected data and make effective predictions. In the recent months, several such models have been developed to understand and predict the transmission of COVID-19 pandemic. See for example [1, 2].

Fractional-order models have been recognized as a powerful mathematical tool to study anomalous behaviors observed in many physical processes with prominent memory and hereditary properties. In general, such properties might not be satisfactorily captured by the integer-order counterpart models. This motivated researchers to consider fractional-order compartmental models for better understanding of dynamics of infectious diseases such as Dengue fever [3, 4], Influenza [5], and recently COVID-19 [6].

In this article, we propose and simulate a system of fractional order differential equations for the dynamics of COVID-19 pandemic. The proposed model uses the time varying transmission rate given in [1] and [7] which incorporates the effects of governmental actions and public perception. Since the focus is to provide adequate study and simulation of the fractional order model, we adopt the two-step generalized time differencing GETD2 scheme developed by Garrappa and Popolizio in [8] to obtain the numerical results. However, to achieve a cost-effective implementation of this scheme, we use the partial fraction decomposition of the global Padé approximants of the two-parameter Mittag-Leffler function [9].

The merit of the proposed fractional model is highlighted by examining the mean square errors between the predicted and reported data in [10]. In addition, an approximation of the optimal fractional order is obtained by minimizing these error. This feature can be used to make better data fitting for subsequent predictions.

The rest of this paper is arranged as follows. Next, the memory-dependent SEIR model is introduced. The computational algorithm is described in Section 3. The simulation results are presented and discussed in Section 4.

## 2. Memory-Dependent COVID-19 Model

Following [1], we assume the total population *N* at any given time to be distributed among the four compartments: susceptible, exposed, infectious, and recovered, labeled by *S, E, I* and *R*, respectively. That is,

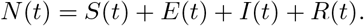

The assumed dynamics among these populations is depicted in Figure 1 and can be describes as follows:

**Figure 1:**
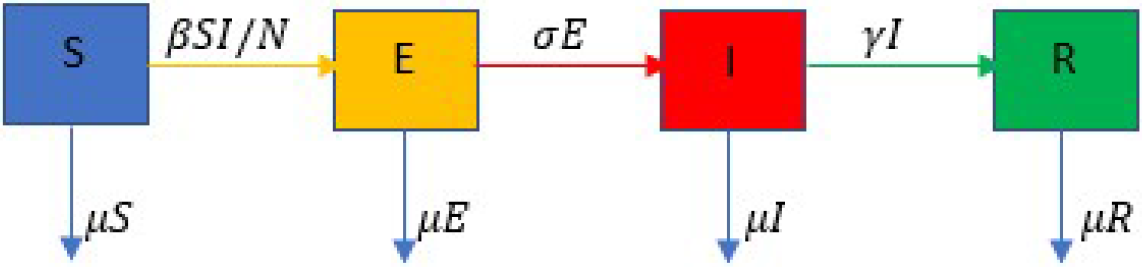
Transfer diagram between the compartments.

- Susceptible individuals enter the exposed compartment at the rate *βI/N*, where *β* is a nonlinear transmission rate.
- Exposed individuals progress to the infectious compartment at the rate *σ*, while the infectious individuals recover from the disease at a rate *γ*.
- The population is assumed to have a natural death rate *µ*.

Furthermore, we consider a variable *P* (*t*) to represent the public perception on the risk of the pandemic. This quantity increases with the number of infections at a rate *φ × γ*, where *φ* is the proportion of severe cases, and diminishes over time at a rate *λ*.

The values *σ*^−1^, *γ*^−1^, and *λ*^−1^ can be interpreted as the mean latent period, mean infectious period, and mean duration of public reaction, respectively. These quantities together with *µ*, and *β* all have units of (time)^−1^.

Based on the above assumptions, we propose the following model for the dynamics of COVID-19 pandemic:

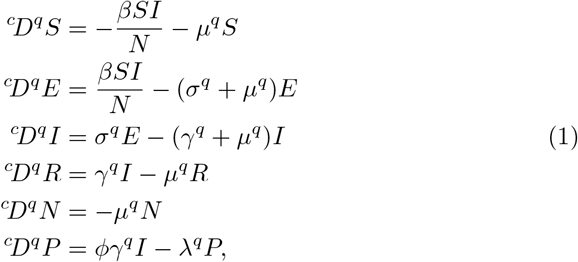

subject to the non-negative initial conditions

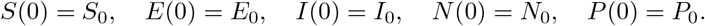

The operator ^*c*^*D*^*q*^ denotes the Caputo fractional derivative of order *q ∈* (0, 1)

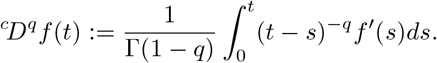

In the special case *q* = 1, the model in (1) reduces to a modification of COVID-19 model introduced in [1].

As observed in the right-hand sided of (1), the *q*th power of the parameters is used as suggested by Diethelm [3] and Angstmann *et al*. [11]. This is necessary to ensure the consistency in units since the left-hand side has the units of (time)^−*q*^.

For the transmission rate *β* in (1), as in [1], we use the formula

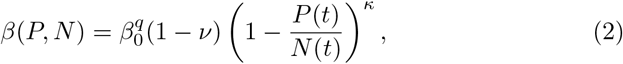

where *ν* is the strength of governmental action, *β*_0_ is the baseline transmission rate, and *κ* is the intensity of individual reaction. Note that the transmission rate decreases as the perception of risk per individual *P/N* and the strength of action *ν* increase. Moreover, the transmission rate reduces to the baseline rate *β*_0_ in the absence of governmental action and lack of any perception of risk.

## 3. Computational Algorithm

Here, we describe briefly the numerical algorithm used to simulate model (1). For this purpose, we consider the matrix form of the system,

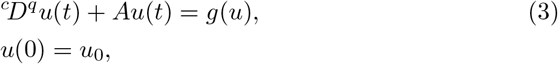

where

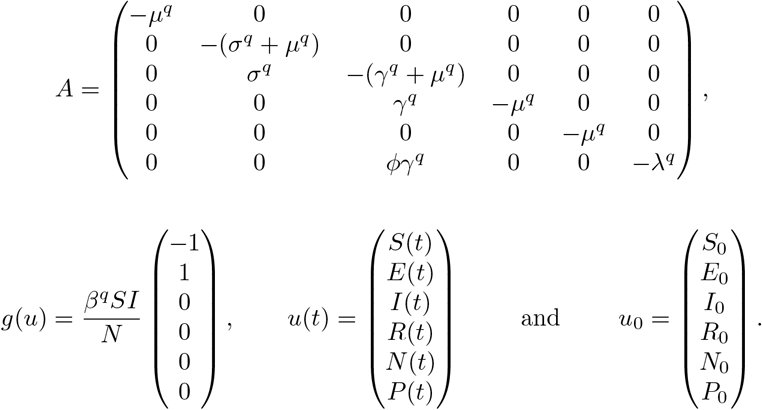

To solve numerically (3) on some interval [0, *T*], we introduce the uniform mesh

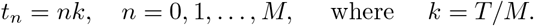

Then, by applying the GETD2 scheme developed in [8], we have the approximation *v*^*n*^ *≈ u*(*t*_*n*_) given by

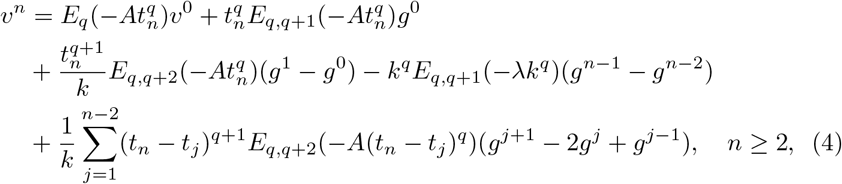

where *g*^*j*^ = *g*(*v*^*j*^). To initialize the scheme we take *v*^*0*^ = *u*_*0*_ and compute *v*^1^ as

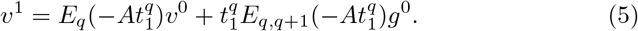

As seen in (4) and (5), the implementation of GETD2 scheme involves matrix-vector products for the Mittag-Leffler function of a matrix.

This implementation can be carried out using the technique discussed in [8]. However, that approach is costly. For efficient implementation of the GETD2 scheme, we use a technique based on the partial fraction decomposition of the global Padé approximant of type (7, 2) developed in [9] for the Mittag-Leffler function.

## 4. Model Simulation

In this section, we present the numerical simulations and demonstrate the effectiveness of model (1) in matching the reported data. For this purpose, we use the parameters given in [1] for China, namely,

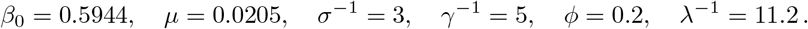

As per the lockdown records of Chinese Administrative Division, we take *ν* = 0 for the first 16 days (Jan 20 – Feb 4) and *ν* = 0.8478 for the period Feb 5 – March 6, together with *κ* = 1117.3. The initial populations are

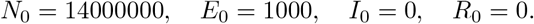

On the other hand, due to the infection-free initial condition, the initial perception *P*_0_ is 0.

In Figures 2 and 3, the simulations highlight the main advantages of adopting the fractional order model (1). With the extra parameter *q*, the model has a better capability of capturing the reported data. In particular, the model performs quite well in capturing the peak and slow decay of the infectious population. This is asserted by examining the mean square errors which also indicate that *q* = 0.955 is a good approximation of the optimal order. The model with optimal fractional order can then be used to make future predictions.

**Figure 2:**
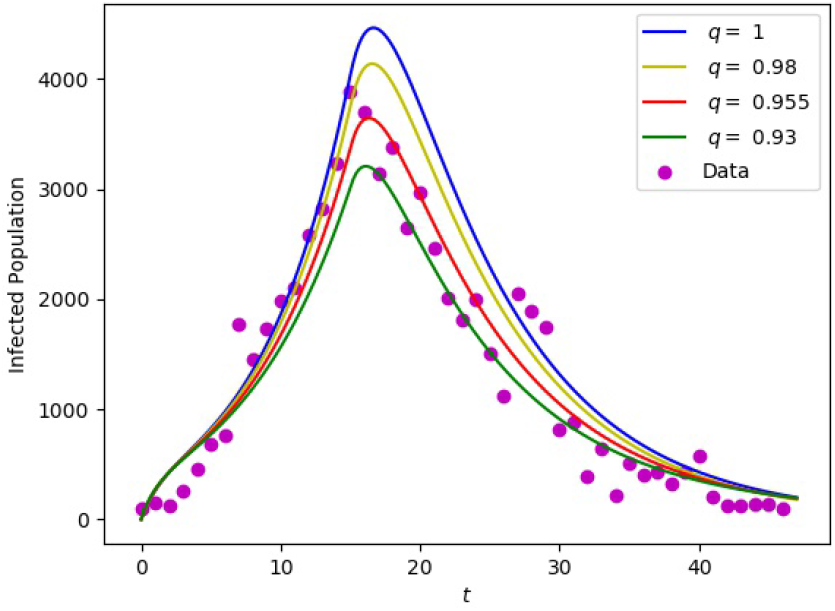
Data matching for different orders *q*.

**Figure 3:**
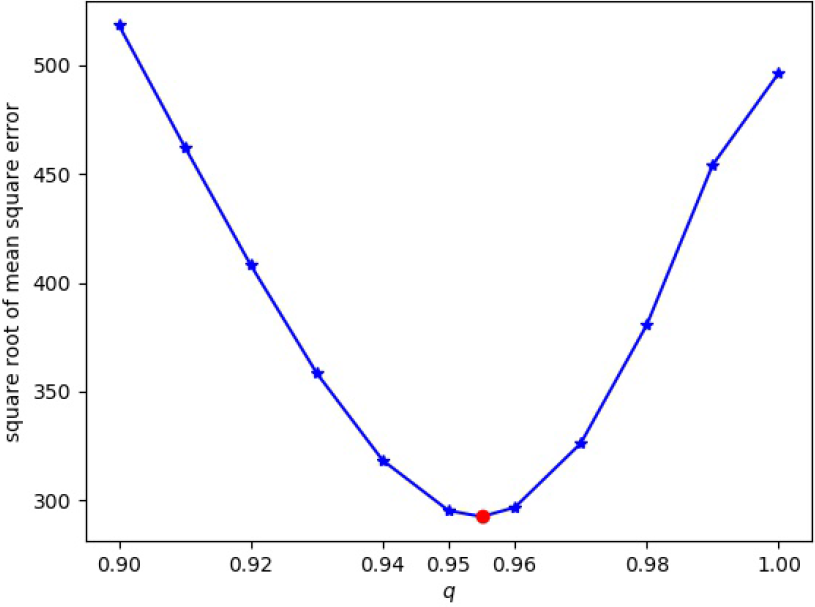
Mean square errors for different orders *q*.

In Figure 4, the transmission rate corresponding to different values of governmental action *ν* and intensity of human behavioral response *κ* are depicted. As observed, for fixed intensity *κ* and fractional order *q*, the transmission rate decreases significantly as the governmental action increases. Similar impact is observed when the intensity *κ* is increased.

**Figure 4:**
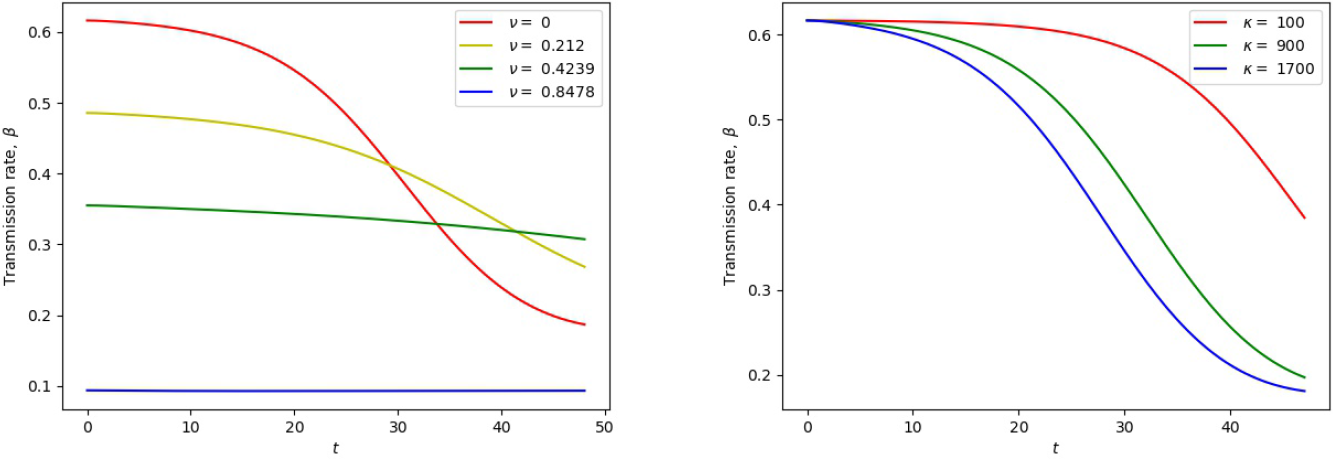
Transmission rate with respect to *ν* and *κ*.

Overall, the obtained results emphasize the robustness of the fractional-order model as a generalization of the integer-order model.

## Conclusion

In this paper, we proposed a nonlinear fractional order model to study the dynamics of COVID-19 pandemic. Using the data of the outbreak in Wuhan, China, the effectiveness of the proposed model in simulating the spreading of the disease is demonstrated. By examining the mean square error of the predicted number of infections in our model and the reported data, we found that the proposed fractional model gives better prediction of the reported data than the integer order model.

## Data Availability

Data

## Acknowledgment

We would like to acknowledge the support provided by King Fahd University of Petroleum & Minerals via the project SB191001.

## Notes

### Competing Interest Statement

The authors have declared no competing interest.

### Funding Statement

No external funding was received

